# Common and Distinct Drug Cue Reactivity Patterns Associated with Cocaine and Heroin: An fMRI Meta-Analysis

**DOI:** 10.1101/2023.10.19.23297268

**Authors:** Jordan M. Dejoie, Nicole Senia, Anna B. Konova, David V. Smith, Dominic S. Fareri

**Author notes:** Corresponding Author: Dr. Dominic Fareri Adelphi University Gordon F. Derner School of Psychology Blodgett Hall, Rm. 212C Garden City, NY 11530. This manuscript is supported in part by funding from National Institute on Mental Health (R15MH122927 to DSF), the National Institute on Drug Abuse (R01DA053282, R01DA054201 to ABK), and the National Institute on Aging (RF1-AG067011 to DVS).

## Abstract

Substance use and substance use disorders represent ongoing major public health crises. Specifically, the use of substances such as cocaine and heroin are responsible for over 50,000 drug related deaths combined annually. We used a comparative meta-analysis procedure to contrast activation patterns associated with cocaine and heroin cue reactivity, which may reflect substance use risk for these substances. PubMed and Google Scholar were searched for studies with within-subject whole brain analyses comparing drug to neutral cues for users of cocaine and heroin published between 1995 and 2022. A total of 18 studies were included, 9 in each subgroup. Voxel-based meta-analyses were performed using seed-based d mapping with permuted subject images (SDM-PSI) for subgroup mean analyses and a contrast meta-regression comparing the two substances. Results from our mean analysis indicated that users of heroin showed more widespread activation in the nucleus accumbens, right inferior and left middle temporal gyrus, right thalamus, and right cerebellum. Cocaine use was associated with recruitment of dorsolateral prefrontal cortex during cue reactivity. Direct comparison of cue reactivity studies in heroin relative to cocaine users revealed greater activation in dopaminergic targets for users of heroin compared to users of cocaine. Differential activation patterns between substances may underlie differences in the clinical characteristics observed in users of cocaine and heroin, including seeking emotional blunting in users of heroin. More consistent research methodology is needed to provide adequate studies for stringent meta-analyses examining common and distinct neural activation patterns across substances and moderation by clinically relevant factors.

## 1. Introduction

The growing prevalence of substance use disorders among the US population, particularly related to illicit substances lacking regulation (i.e., opiates, psychostimulants), thus represents a significant threat to public health. Though the variety of illicit drugs available for use has increased over time, stimulants (e.g., cocaine) and opioids (e.g., heroin, fentanyl, prescription medications) remain two of the most lethal classes of illicit drugs (Degenhardt et al.,2013). In fact, opioids and stimulants in particular account for almost 81,000 and 15,000 deaths annually, together representing approximately 80% of all substance related deaths (National Institute on Drug Abuse, 2022; CDC, 2023). Given the high rates of mortality associated with cocaine and heroin use relative to other substances (Schneider et al., 2021; Peacock et al., 2021; Lewer et al., 2022; NIDA, 2022; CDC, 2023; Darke et al., 2023), we selected these substances as targets for the present study. These substances are also representative of the broader opioid and stimulant drug classes, whose use (and co-use) has surged nationally in what has been labeled as the “twin opioid-stimulant epidemics”. Lastly, although within the broader class of stimulant and opioid drugs, others have received considerable recent attention (e.g., fentanyl), cocaine and heroin remain the most well studied in fMRI studies of cue reactivity.

Nevertheless, differences in usage patterns between these classes of drugs have also been well-documented. For example, heroin use is often associated with a more consistent, chronic and high rate of use whereas cocaine tends to involve more episodic, intermittent, and moderate use (Hser et al., 2008a; Hser et al., 2008b; Sartor et al., 2014). Users of heroin also tend to begin use at a younger age and as a result have a longer duration of lifetime use than people who use cocaine (Hser et al., 2008a; Hser et al., 2008b). Onset of heroin, and opioid use often occurs in conjunction with traumatic events, and with a desire to numb both physical and emotional pain (Purdue et al., 2024; Shand et al., 2011), while users of cocaine may demonstrate higher levels of impulsivity and impaired inhibition, resulting in higher rates of risk-taking and reward-seeking behaviors such as sexual promiscuity compared to people who use heroin (Lejuez et al., 2005; Bornovalova et al., 2005; Mitchell & Potenza, 2014; Bjork et al., 2022; Vassileva et al., 2014; Ahn & Vassileva, 2016). As these behavioral differences do not appear to depend on acute drug effects, it is possible that they may reflect differences in susceptibility between those who use cocaine versus heroin.

Interestingly, whether one is in a social or non-social environment appears to impact drug choice (Caprioli et al., 2009; De Pirro et al., 2018). In both animal and human models, heroin use tends to be more common within non-social contexts, whereas cocaine use is often preferred in social environments (Caprioli et al., 2009; Badiani et al., 2011; Badiani et al., 2019). Such contextual preferences also amplify the respective effects of each substance (De Pirro et al., 2018). In sum, these differences suggest that in addition to shared mechanisms, there may also be divergent mechanisms that are important to identify to better understand patterns of usage of cocaine and heroin. Identifying divergent mechanisms (i.e., pinpointing behavioral and neurobiological risk-factors for the development of a particular SUD, developing drug-specific treatment interventions) is imperative given the rapidly increasing rates of use and associated harm caused by these illicit substances.

A common component of substance use disorders that is closely tied to reuse and relapse is drug craving (i.e., the strong wanting or desire for a substance; Vafale & Kober, 2022). Characterizing the ways in which the brain differentially responds to craving in people who use cocaine and heroin may provide an important way to assess shared and distinct vulnerability mechanisms and may have implications for developing more targeted interventions to prevent their use. Cue-reactivity paradigms are reliably related to subjective experiences of drug cravings and are thus widely used, well– supported laboratory assessments in samples of individuals using substances (Drummond, 2000; Carter & Tiffany, 1999), and have been used previously to reveal similarities and differences between individuals who use stimulants, opioids, or both (e.g., Hochheimer et al., 2023). In fact, a recent study found that when comparing cue induced and tonic craving between opioids and stimulant users in treatment, opiate cues consistently resulted in greater reports of craving (Hochheimer et al., 2023). Typically, such paradigms involve exposure to a drug cue or set of drug cues (e.g., images of a drug or associated paraphernalia, auditory drug cues via stories or sounds) and the subsequent measurement of self-reported craving or physiological responses to these cues (e.g., psychophysiological signals, patterns of neural activation; Carter & Tiffany, 1999; Drummond, 2000; Courtney et al., 2016).

Task-based fMRI studies implementing cue-reactivity paradigms have been extensively used to characterize the neural circuitry of substance use disorders, including for identifying common and specific patterns associated with use of licit and illicit substances (e.g., Chase et al., 2011; Kuhn & Gallinat, 2011; Zilberman et al., 2019). However, to our knowledge, there have been no systematic comparisons that focus specifically on cocaine versus heroin drug cue reactivity, although such comparisons would be highly relevant given ongoing public health concerns associated with these substances and clinically relevant differences in their usage. Studies have revealed that targets of dopaminergic input such as the striatum, prefrontal cortex, and thalamus are recruited during cocaine use (Haber & Knuston, 2010; Wang, Smith & Delgado, 2016; Bartra et al., 2013; Huang et al., 2018) and play a key role in craving and sustaining the cycle of addiction, consistent with rodent models (Jentsch & Taylor, 1999; Goldstein & Volkow, 2002; Everitt & Robbins, 2005; Huang et al., 2018). Similarly, heroin recruits dopaminergic targets such as the dlPFC, insula, and orbitofrontal cortex (OFC; Liu et al., 2021) it also seems to target other systems including the glutamatergic system (Gheraldini et al., 2015) which conceptualized as the basis for heroin’s analgesic effect.

Further, prior meta-analyses studying users of these substances have largely utilized broad inclusion criteria, or examined outcomes other than functional activation (e.g., gray matter, structural differences; Klugah-Brown et al., 2021; Pando-Nuande et al., 2021; Hill-Bowen et al., 2022; Long et al., 2022). Although, studies have begun to identify common neural circuits implicated in cocaine and heroin use, there is no existing meta-analytic synthesis of the extant literature to quantitatively evaluate similarities and differences in the functional neural underpinnings of cocaine versus heroin use. These methodological differences impact the ability to characterize similarities and differences in cue reactivity between users of each substance. Additionally, an important factor to consider is whether similarities/differences in neural recruitment during cue reactivity across users of cocaine and heroin vary as a function of individual differences in length of drug use/chronicity. Thus, a more stringent synthesis regarding the neural circuits implicated in cocaine and heroin usage is needed.

To address these gaps, we conducted a meta-analysis of fMRI studies aimed at comparing activation patterns during cue-reactivity tasks for drug relative to neutral cues in people who use cocaine and people who use heroin. We applied strict inclusion criteria for our meta-analysis such as utilizing studies implementing within subjects, whole brain analyses of drug vs neutral cues. We expected that studies of both people who use cocaine and people who use heroin will show significant engagement of reward-related and dopaminergic circuits during cue-reactivity tasks, given the well-documented recruitment of these circuits by both substances. We also expected that, people who use cocaine will exhibit greater activation in corticostriatal circuits than people who use heroin, given possible differences in levels of impulsivity, risk taking, and impaired inhibition observed in cocaine relative to heroin. We also explored the impact of length of use on neural correlates of cue reactivity, with the expectation that length of use would be negatively associated with activation in dopaminergic regions including the prefrontal cortex and regions associated with reward processing such as the striatum.

## 2. Method

### 2.1 Literature search and study selection

A literature search was conducted via PubMed and Google Scholar (up to April 2022) to identify applicable studies. Keywords used were (‘cocaine’, ‘heroin’, ‘substance use’, ‘cue-reactivity’, ‘fMRI’). Search strings used were (‘cocaine fMRI cue-reactivity’, ‘heroin fMRI cue-reactivity’) along with other combinations of the keywords. All reference lists from identified studies were checked manually for additional applicable studies. In the current meta-analysis, studies were included if (1) analyses were conducted via fMRI; (2) participants were medically healthy aside from their substance use adults over the age of 18; (3) participants were either abstaining from or currently engaging in drug use; (4) the study included a substance vs. neutral condition whole-brain contrast within the substance using group; (5) the study utilized a cue-reactivity paradigm; (6) reported coordinates in either Montreal Neurological Institute (MNI) or Talairach and Tournoux space. Studies were excluded if (1) they lacked a substance vs. neutral whole brain contrast analysis; (2) included PET data; (3) there was insufficient reporting of fMRI data or findings; (4) they did not report coordinates from whole-brain analyses; (5) neither people who use cocaine nor heroin were included in the study; (6) analyses were not separated according to drug type.

Studies were selected based on the above inclusion and exclusion criteria and were reviewed by all authors (JD, NS, DF). The corresponding author made final decisions regarding inclusion/exclusion conflicts. Study quality was assessed using a recently proposed checklist for cue-reactivity studies (Ekhtiari et al., 2022). Given the recency of this publication compared to the dates of publication for our studies we set our threshold for quality at 50% of criterion met within a category for at least 5 of the 10 proposed categories. All studies included in the current study exceeded this threshold meeting criterion for at least 7 of the 10 proposed categories. Figure 1 depicts the process of literature search and study selection. 1025 studies were first identified via the literature search; any duplicates from searches were removed. Next, 949 studies were excluded based on title/abstract review. The remaining 76 full text articles were assessed for eligibility. Exclusion of studies occurred for a variety of reasons including: 1) participant groups were composed of people who use opiates, in general, as opposed to solely people who use heroin; 2) only between-group analyses were conducted comparing people who use substances to healthy controls––no within-subjects analyses examining responses to drug versus neutral cues; 3) lack of reporting of whole brain analyses, a focus on only connectivity or solely ROI analyses; 4) Lastly, an implementation of experimental tasks other than a drug versus neutral cue task (i.e., stroop word task, drug versus food cues, etc.). Of the 76 full-text studies, 58 studies were excluded based on the above criteria, leaving 18 studies remaining (total N = 447) Each subgroup––cocaine and heroin––was composed of 9 studies (N_cocaine_=273; N_heroin_=174) that were included in this meta-analysis (see Table 1). Group differences in sample characteristics, compared using independent samples t-tests, across included studies are presented in Table 3.

**Figure 1.**
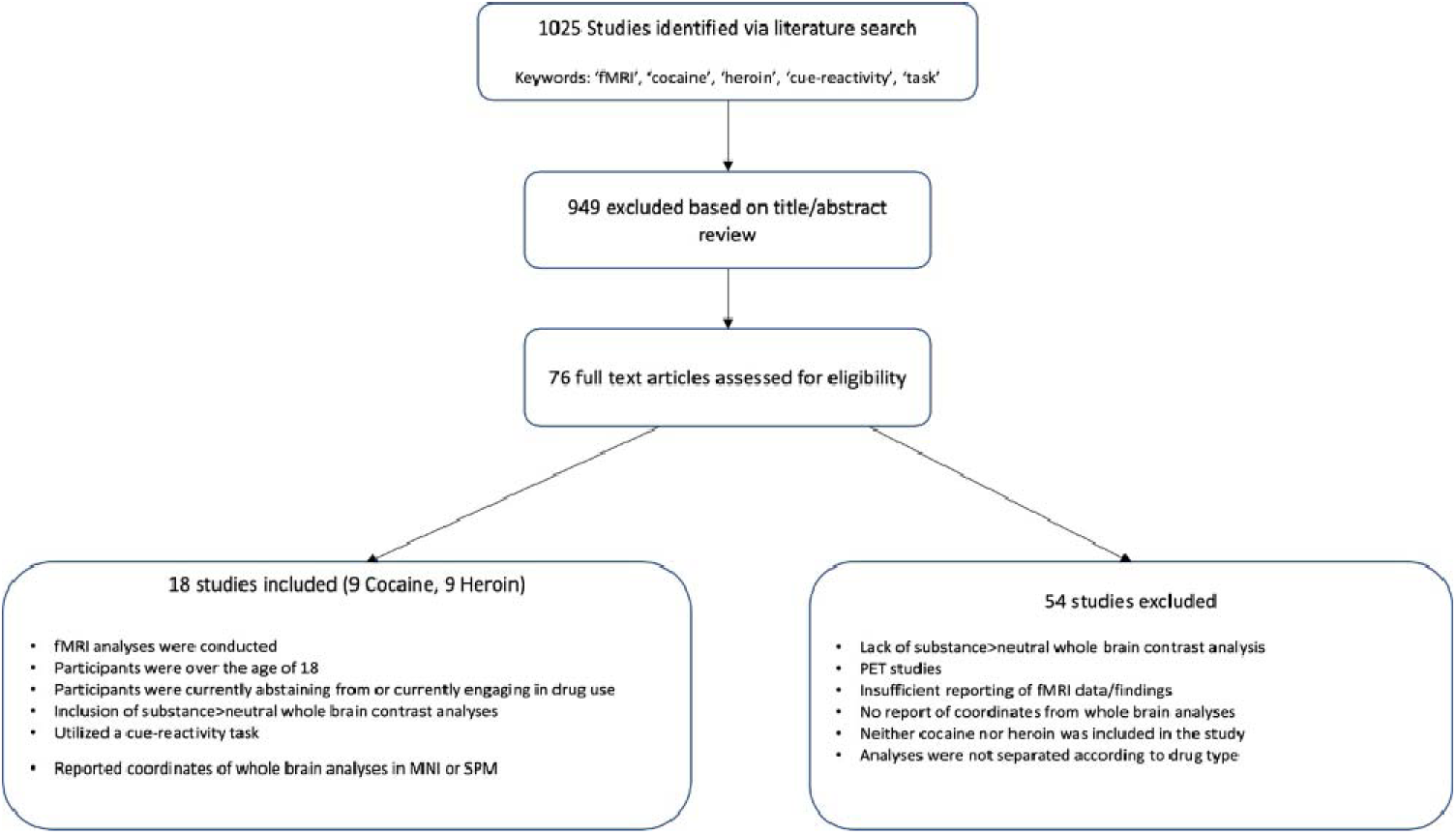
Flowchart PRISMA diagram outlining the process of literature search and study selection.

**Table 1.** Characteristics of 18 fMRI studies included in the contrast meta-analysis, sub-group mean analyses, and the 15 included in exploratory meta-regression.

| Author | Year | N | Task | Drug | Space | Included in Meta-Regression |
| --- | --- | --- | --- | --- | --- | --- |
| Duncan, E., Boshoven, W. Harenski, K. et al. | 2007 | 10 | Script guided mental imagery | C | MNI | Y |
| Elton, A., Smitherman, S., Young, J. et al. | 2014 | 20 | Script guided mental imagery | C | MNI | Y |
| Goldstein, R., Tomasi, D., Alla-Klein, N. , et al. | 2009 | 15 | Word-Cue Task | C | TAL | Y |
| Hanlon, C.A., Dowdle, L.T., Gibson, N.B. et al. | 2018 | 55 | Image-Cue Task | C | MNI | N |
| Hassani-Abharian, P., Ganjgahi, H., Tabatabaei-Jafari, H., et al | 2015 | 20 | CRET task | H | MNI | Y |
| Kaag, AM, Reneman, L., Homberg, J., et al. | 2018 | 59 | Image-Cue Task | C | MNI | Y |
| Li, Q., Wang, Y., Zhang, Y., et al. | 2012 | 24 | Image-Cue Task | H | MNI | Y |
| Li, Q., Wang, Y., Zhang, Y., et al. | 2013 | 19 | Image-Cue Task | H | MNI | Y |
| Mei, W., Zhang, J.X., & Xiao, Z | 2010 | 12 | Image-Cue Task | H | MNI | Y |
| Prisciandaro, J.J., Myrick, H., Henderson, S. McRae-Clark, A.L., Santa Ana, E.J. et al. | 2013 | 25 | Image-Cue Task | C | MNI | Y |
| Prisciandaro, J.J., Myrick, H., Henderson, S., McRae-Clarck, A.L., & Brady, K.T. | 2013 | 30 | Image-Cue Task | C | MNI | N |
| Prisciandaro, J.J., McRae-Clark, A.L., Myrick, H., et al. | 2014 | 38 | Image-Cue Task | C | MNI | Y |
| Walter, M., Denier, N., Gerber, H., et al. | 2014 | 27 | Image-Cue Task | H | MNI | Y |
| Wang, W., Li, Q., Wang, Y. et al. | 2011 | 14 | Image-Cue Task | H | MNI | Y |
| Wang, A-L. Elman, I., Lowen, S.B., et al. | 2015 | 32 | Event Related and Image Cue Tasks | H | TAL | N |
| Xiao, Z., Lee, T., Zhang, J.X., et al. | 2005 | 14 | Image-Cue Task | H | TAL | Y |
| Zhang, S., Zhornitsky, S., Wang, W. et al. | 2020 | 52 | Image-Cue Task | C | MNI | Y |
| Zijlstra, F., Veltman, D.J., Booji, J., et al. | 2009 | 12 | Image-Cue Task | H | MNI | Y |

### 2.2 Seed based d mapping

Seed-based *d* mapping (SDM) SDM-PSI v. 6.22 (formerly *Signed Differential Mapping)* is a specific statistical method used for meta-analyzing structural and/or functional differences in the brain across studies using a variety of different neuroimaging techniques including fMRI, VBM, DTI, or PET (Albajes-Eizagirre et al, 2019a). The SDM method consists of a series of steps. To begin, coordinates of identified cluster peaks from individual studies are entered. Next, during pre-processing, the upper and lower bounds of possible effect sizes (Hedges g) and variances are calculated for each study. A mean-analysis is then performed where maximum-likelihood estimation (MLE) and MetaNSUE are implemented to estimate the most likely effect size for each included study and its standard error (Albajes-Eizagirre et al., 2019a). MetaNSUE is a meta-analytic method that decreases bias and permits inclusion of non-statistically significant unreported effects (NSUEs) (Albajes-Eizagirre et al., 2019b). MetaNSUE involves multiple imputations that are subsequently created by adding noise to the estimations within the previously calculated bounds (Albajes-Eizagirre et al., 2019b). Each of the imputed data sets are then meta-analyzed, and Rubin’s rules (a pooling of parameter estimates in an imputed data set to provide confidence intervals and p-values) are applied to combine the imputed meta-analyzed datasets. Lastly, a subject-based permutation test is conducted in which the maximum statistics, (i.e., the largest z-value reflecting the peak of activation across the whole brain in each study), from the combined meta-analysis images are saved (Albajes-Eizagirre et al., 2019b). A family-wise error rate correction (FWER) for multiple comparisons is then performed using the distribution of the maximum statistic.

### 2.3 Meta-Analysis Procedure

Coordinates of peak activation and additional pertinent information (e.g., statistical values [*p*-value, *t*-value, *z*-value], statistical thresholds, and sample sizes) for each within-group drug cue > neutral contrast from the included studies were collected and converted into text files. Any *p-*values or *z-*values reported in the studies were converted to *t*-values using the SDM online converter. We then conducted a voxel-based meta-analysis using SDM-PSI v. 6.22 for all studies included, irrespective of drug type, and for each subgroup. We also conducted a subsequent linear model. The same parameters were utilized for all analyses and are outlined as follows. Pre-preprocessing for each group was conducted within a gray matter mask using the default settings within SDM-PSI: a 20-mm anisotropic full width half maximum (FWHM) kernel and 2-mm voxel size (Albajes-Eizagirre et al., 2019a). The mean analysis and permutation tests were then performed within each group of studies separately (cocaine, heroin), with the number of imputations set to 50 and the number of permutations set to 5000. The results of the meta-analyses of the subgroups were then corrected using the FWER correction with 5000 permutations and subsequently thresholded at *p* < .05.

### 2.4 Contrast Analysis

We conducted a linear regression to compare the two subgroups, Heroin > Cocaine and Cocaine > Heroin. The inter-study heterogeneity of each cluster surviving correction was measured by the *I^2^ index,* which represents the proportion of total variation due to study heterogeneity (Higgins and Thompson, 2002). *I^2^* > 50% commonly indicates considerable heterogeneity. The maps resulting from the contrast analyses were also corrected to a level of *p* < .005 TFCE-FWER, with a cluster extent threshold of ≥ 10 voxels. Subsequent bias tests (e.g., funnel plots, I^2^ index, metabias test, and excess significance tests) were conducted for this analysis using the SDM software. The I^2^ value is generated during the extraction of the peaks and represents the percentage of heterogeneity across the data and studies included in the meta-analysis (Albajes-Eizagirre et al., 2019c). The funnel plot test identified if there was asymmetry in the plot (i.e., whether there were larger effect sizes in smaller studies). Last, the excess significance test indicates if there was publication bias (i.e., whether studies are only published if the results are significant; Albajes-Eizagirre et al., 2019c).

### 2.5 Exploratory Meta-Regression

We also tested whether length of use moderated activation by incorporating length of use and a substance type x length of use interaction into the model. Due to limited data in the included studies regarding demographic and clinical features, no other features were included as meta-regressors. The inter-study heterogeneity of each cluster surviving correction was measured by the *I^2^ index,* which represents the proportion of total variation due to study heterogeneity (Higgins and Thompson, 2002). *I^2^* > 50% commonly indicates considerable heterogeneity. The maps resulting from the contrast analyses were also corrected with *p* < .005 TFCE-FWER, with a cluster extent threshold of ≥ 10 voxels. Nilearn was used to visualize results from all analyses (Abraham et al., 2014).

## 3. Results

### 3.1 Clinical and Demographic Differences

Assessment of group differences in age and length of use revealed a significant difference in age with cocaine participants being significantly older but no difference in length of use (See Table 3 for full description of study sample characteristics). Further, all studies excluded participants with a co-occurring SUD outside the drug of interest in the study; however, most studies permitted recreational tobacco and alcohol use. Four studies permitted inclusion of participants with mood disorders but did not specify whether any participants met criteria. Abstinence was reported variably across studies, making it difficult to directly compare and determine participants’ length of abstinence at the time of participation. When evaluating abstinence based on whether participants were abstinent at data collection (yes/no), participants across all studies were largely abstinent. Overall, participants were largely biological males. In fact, 8 of the 16 studies were an all-male sample. Of the remaining 8 mixed-sex studies, 7 of them were still composed of a majority of male participants (See Tables 2 & 3). In general, cocaine studies appeared to have more mixed-sex samples, though still largely male dominant. Treatment history was evaluated based on whether participants were undergoing medication assisted treatment at the time of data collection. Across all studies, participants were not undergoing treatment except those in two of the heroin studies.

**Table 2.** Clinical characteristics of heroin and cocaine subgroups. Abstinence reflects whether participants were free from substances at the time of data collection.

| **Study** | **Drug** | **Abstinent (Y/N)** | **Co-Use** | **Sex** | **Mean Age (years)** | **Length of Use (mos)** |
| --- | --- | --- | --- | --- | --- | --- |
| Duncan, E., Boshoven, W. Harenski, K. et al. | C | Y | No | Male | 43.6 | 15.9 |
| Elton, A., Smitherman, S., Young, J. et al. | C | N | No | Male | 40 | 176.4 |
| Goldstein, R., Tomasi, D., Alla-Klein, N. , et al. | C | N | Alcohol, Tobacco, Marijuana | 12 M/ 3 F | 43.6 | 188.4 |
| Hanlon, C.A., Dowdle, L.T., Gibson, N.B, et al. | C | Y | Nicotine | 38 M/17 F | 42.7 | NA |
| Hassani-Abharian, P., Ganjgahi, H., Tabatabaei-Jafari, H., et al | H | N | No | Male | 31.9 | 36.24 |
| Kaag, AM, Reneman, L., Homberg, J., et al. | C | N | No other opiates | Male | 31.4 | 72 |
| Li, Q., Wang, Y., Zhang, Y., et al. | H | Y | No other opiates | Male | 32.8 | 78.6 |
| Li, Q., Wang, Y., Zhang, Y., et al. | H | Y | No | Male | 32.2 | 80.5 |
| Mei, W., Zhang, J.X., & Xiao, Z | H | Y | No | 12 M/1 F | 33.5 | 54 |
| Prisciandaro, J.J., Myrick, H., Henderson, S. McRae-Clark, A.L., Santa Ana, E.J. et al. | C | Y | Tobacco, Nicotine | Male | 48.8 | 215.6 |
| Prisciandaro, J.J., Myrick, H., Henderson, S., McRae-Clarck, A.L., & Brady, K.T. | C | Y | Marijuana, Alcohol, Nicotine | 24 M/6 F | 48.4/31.2 | NA |
| Prisciandaro, J.J., McRae-Clark, A.L., Myrick, H., et al. | C | Y | No | 33 M/5 F | 47.7/43.96 | 213.5 |
| Walter, M., Denier, N., Gerber, H., et al. | H | Y | No | 19 M/8 F | 41.1 | 253 |
| Wang, W., Li, Q., Wang, Y. et al. | H | Y | No | Male | 36.1 | 94.8 |
| Wang, A-L. Elman, I., Lowen, S.B., et al. | H | Y | No | 17 M/15 F | 29.19 | NA |
| Xiao, Z., Lee, T., Zhang, J.X., et al. | H | Y | No | Male | 33.2 | 85.2 |
| Zhang, S., Zhornitsky, S., Wang, W. et al. | C | Y | No | 42 M/10 F | 46.7/41.0 | 189.4 |
| Zijlstra, F., Veltman, D.J., Booij, J., et al. | H | Y | No | Male | 42.8 | 192 |

**Table 3.** Group differences in age and length of use across subgroups.

|  | Heroin | Cocaine | T | p -value |
| --- | --- | --- | --- | --- |
| Age (m, SD, range) | 34.75, 4.48, 29.2-42.8 | 42.18, 4.88, 31-48.8 | 3.36 | p < .01 |
| Length of Use (m, SD, range) | 109.29, 74.11, 36.2-253 | 154.31, 78.3, 15.9-215.6 | 1.14 | p > .05 |
| Abstinent at time of study | 3/9 | 1/9 | -- | -- |
| Co-Occurring SUDS | 0/9 | 0/9 | -- | -- |
| Treatment (Medication assisted at time of study) | 2/9 | 0/9 | -- | -- |
| Biological Sex (% Male) | 86% (174 total) | 87% (304 total) | -- | -- |

### 3.2 Aggregate Meta-Analysis

Five clusters/peaks were identified as statistically significant across all studies for drug cues > neutral cues following the general meta-analysis in SDM (see Table 4 and Figure 2). Regions within these clusters (peak x, y, z coordinates reported in MNI space) include the (1) right posterior cingulate (6, -44, 30), (2) right inferior frontal gyrus (48, 12, 32) (3) left inferior frontal gyrus (-50, 10, 28), (4) left superior frontal gyrus (-4, 54, 16), and (5) left inferior temporal gyrus (-46, -68, -8). One peak, the left inferior temporal gyrus, had an *I^2^* value that exceeded 40% (I^2^ = 52%), suggesting that the between-study variance was low with most variance attributed to within study sampling error, and therefore, the metabias test for this peak was not interpreted. The remaining peaks did not have *I^2^*values that exceeded 25% and therefore, metabias tests were interpreted. Metabias tests on the remaining peaks revealed an additional cluster, the left inferior frontal gyrus that demonstrated bias/heterogeneity (p >.05). The *T^2^* value was reviewed for this peak and revealed low dispersion (*T^2^* = .01). All other findings were supported as Metabias test results were non-significant (all *p’s* > 0.2).

**Table 4.** Location of neural activations for aggregate meta-analysis of Heroin and Cocaine studies for drug > neutral cues. Peaks organized by decreasing voxel size.

| ID | Region | X | Y | Z | Voxels | P | SDM - Z |
| --- | --- | --- | --- | --- | --- | --- | --- |
| 1 | Right posterior cingulate gyrus, BA 23 | 6 | -44 | 30 | 1817 | <.001 | 5.915 |
| 2 | Right inferior frontal gyrus, opercular part, BA 44 | 48 | 12 | 32 | 620 | <.001 | 7.222 |
| 3 | Left inferior frontal gyrus, opercular part, BA 44 | -50 | 10 | 28 | 185 | .002 | 6.058 |
| 4 | Left superior frontal gyrus, medial, BA 10 | -4 | 54 | 16 | 223 | .01 | 4.970 |
| 5 | Left inferior temporal gyrus. BA 37 | -46 | -68 | -8 | 75 | .02 | 4.932 |

**Figure 2.**
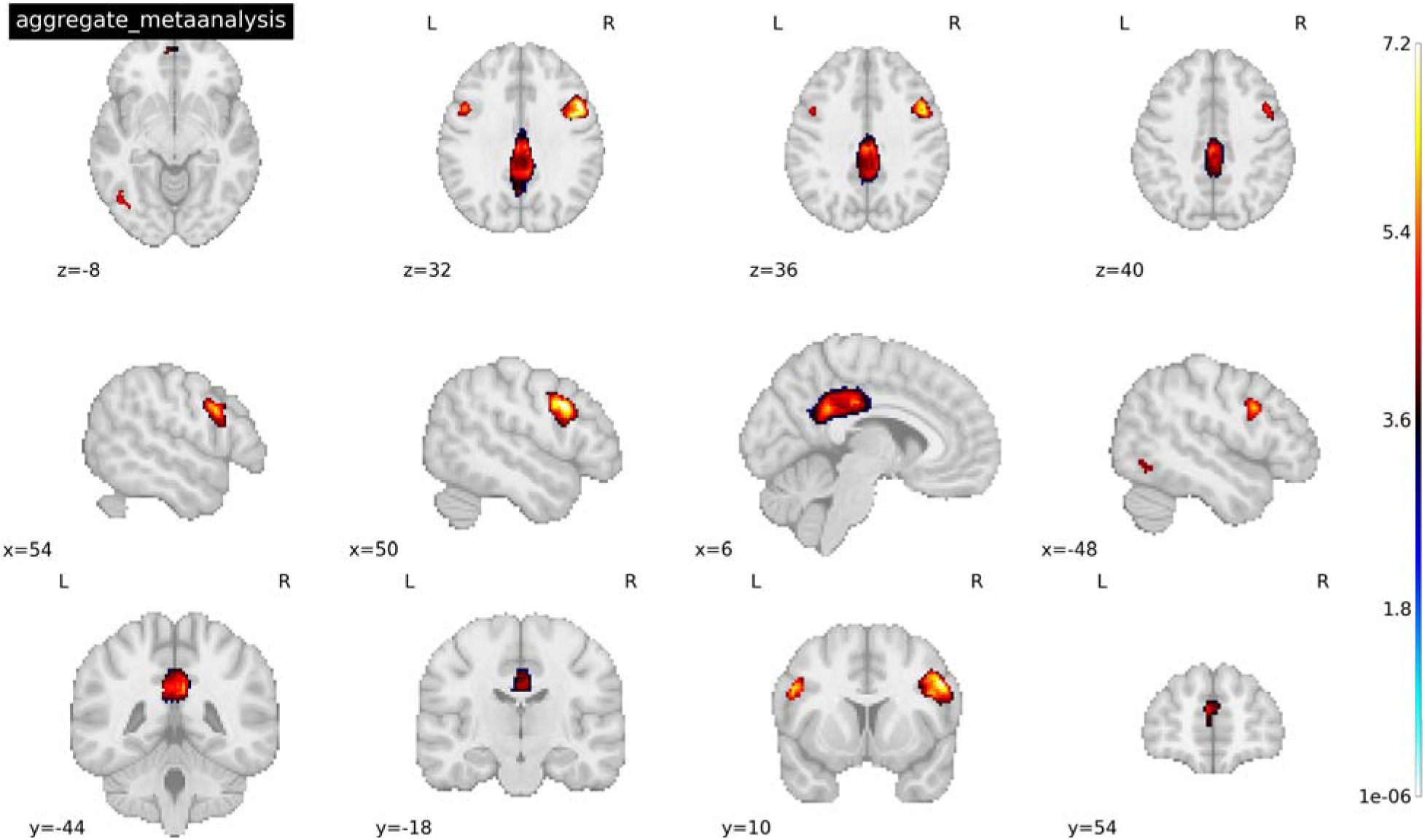
Activated peaks associated with the aggregate mean analysis of 18 cocaine and heroin cue-reactivity studies for drug > neutral cues. Threshold was set at p<.005 TFCE-FWER and cluster size ≥ 10 voxels. Coordinates reported in MNI space. See Table 4 for a full list of activated peaks.

### 3.3 Meta-Analysis of Subgroups

Fourteen clusters/peaks were identified as statistically significant in the Heroin subgroup for drug cues > neutral cues following the general meta-analysis in SDM (see Figure 3). Regions within these clusters (peak x, y, z coordinates reported in MNI space) include the (1) left medial cingulate (-2,30,32), (2) right superior (6, 48, 42) and inferior (48,14,32) frontal gyri, (3) right anterior thalamus (6,48,42), (4) left amygdala (-28,-2,-22), (5) left inferior temporal gyrus (-48,-66,-6), and (6) right cerebellum (24,-74,-44). For a full list of significantly activated regions and their respective coordinates, see Table 5. Only one cluster/peak––right precentral gyrus/inferior frontal gyrus (46, 4, 30) was identified as statistically significant in the cocaine subgroup for drug cues > neutral cues following the general meta-analysis (see Figure 4). Considering the small number of studies (less than ten) that fell within each subgroup, bias test results were conducted but not interpreted (SDM-PSI v. 6.22). However, based on the results of the aggregate meta-analysis results, the left inferior temporal gyrus and left inferior frontal gyrus peaks revealed in the heroin only meta-analysis may be biased.

**Figure 3.**
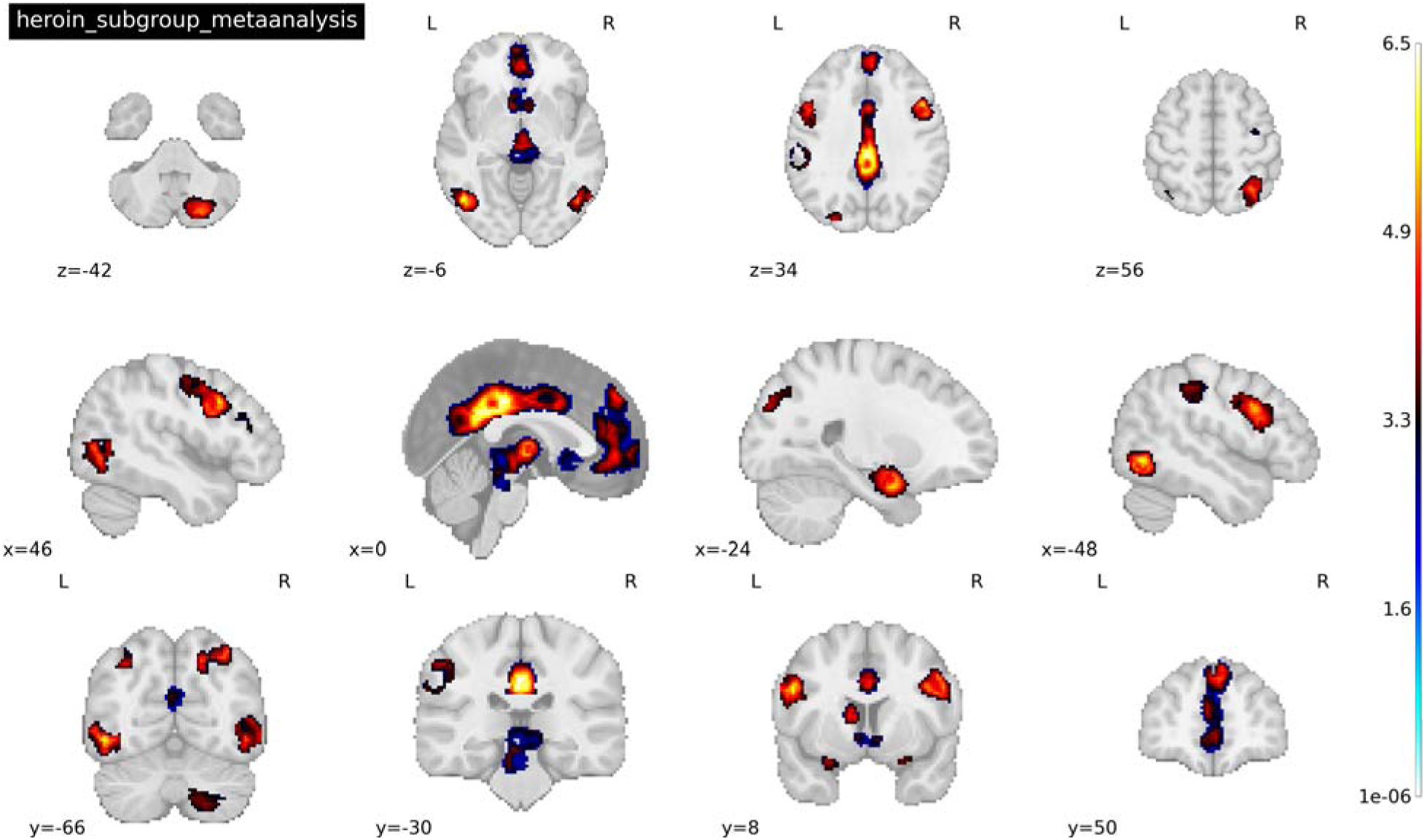
Activated peaks associated with the mean analysis of 9 heroin cue-reactivity studies where drug > neutral cue. Threshold was set at p<.005 TFCE-FWER and cluster size ≥ 10 voxels. Coordinates reported in MNI space. See Table 4 for a full list of activated peaks.

**Figure 4.**
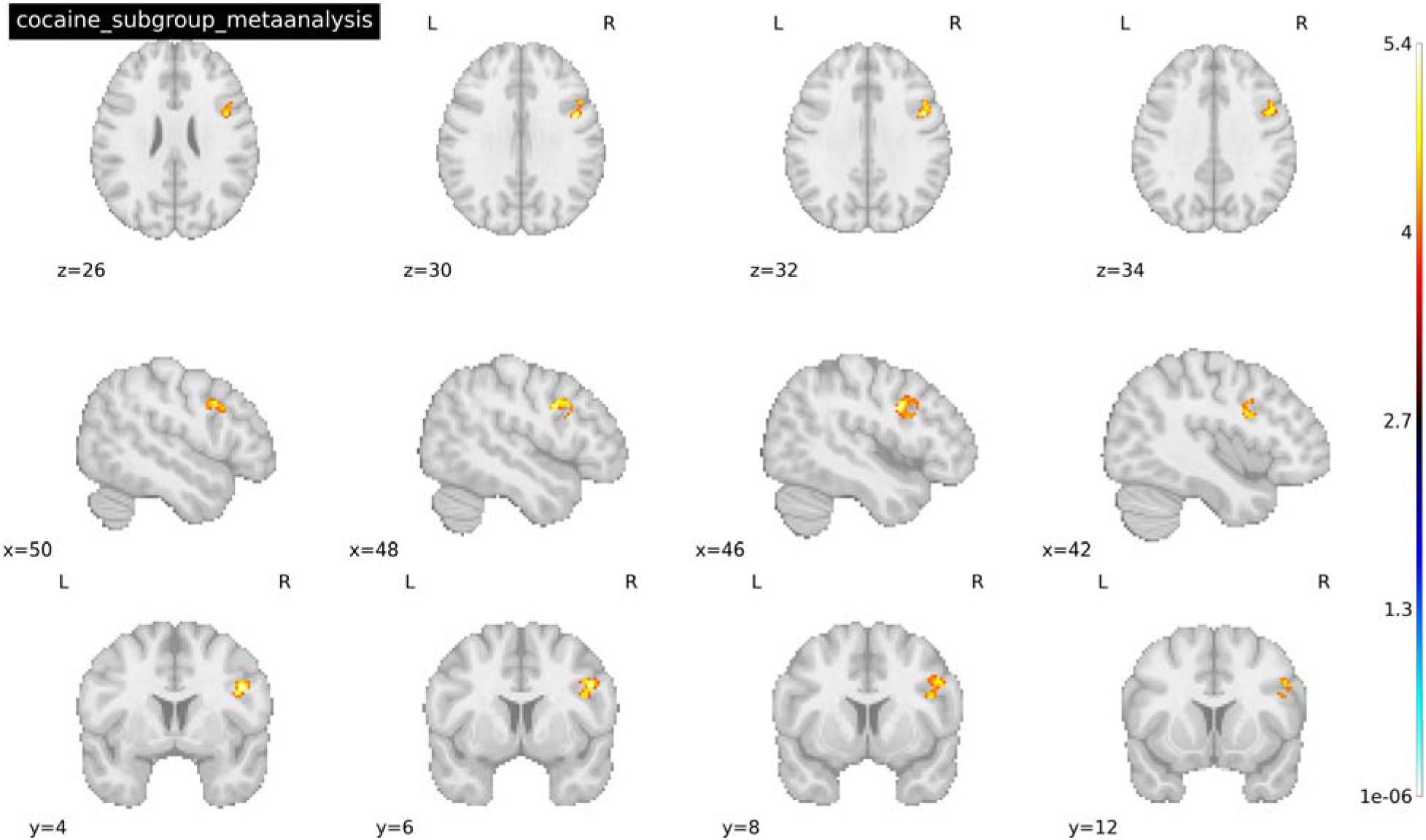
Activated peaks associated with the mean analysis of 9 cocaine cue reactivity studies where drug cue < neutral cue. Threshold was set at p<.005 TFCE-FWER and cluster size ≥ 10 voxels. Coordinates reported in MNI space. Image shows precentral gyrus.

**Table 5.**
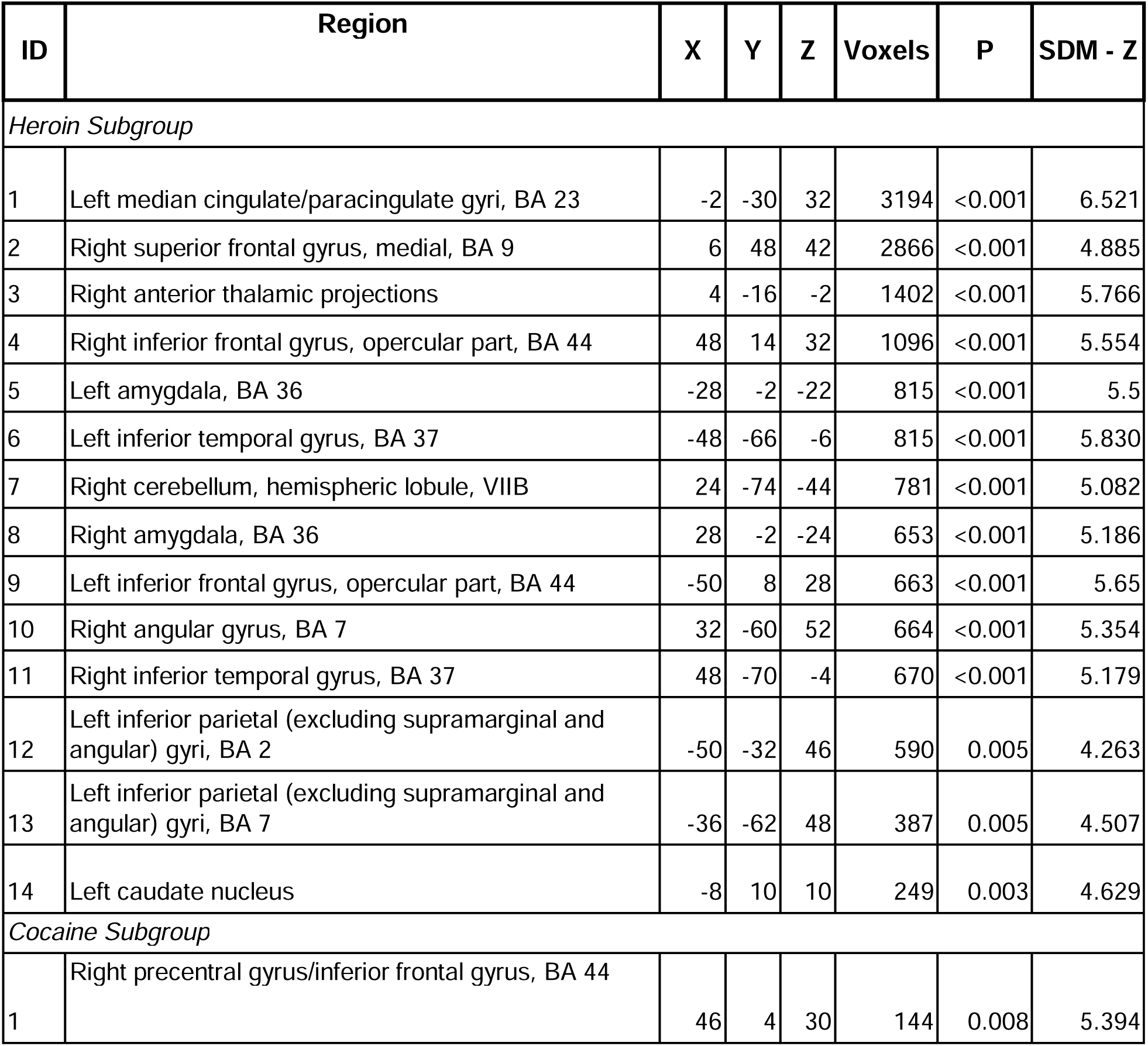
Location of neural activations for mean analysis Heroin and Cocaine subgroups for drug > neutral cues. Peaks organized by decreasing voxel size.

### 3.4 Contrast analysis

Our primary interest was in examining differential recruitment of neural circuits at the whole brain level between the cocaine and heroin subgroups in response to drug relative to neutral cues. We performed a contrast analysis to investigate this question. We observed increased activation in the Heroin > Cocaine contrast in several regions (see Figure 5) including: (1) right inferior (48,-74,-4) and superior (30, -68,50) parietal gyrus; (2) thalamus (6,-14,2); (3) caudate nucleus (-10,10,10); (4) right superior frontal gyrus (6,48,40); and (5) right cerebellum (20,-80,-40). For a full list of significantly activated regions and their respective coordinates see Table 6. Low between-study heterogeneity was found for each significant peak (*I^2^* = 0.74 - 32.2%). Funnel plots were symmetric, suggesting that none of the results were a function of a small subset of included studies or by studies with a small sample size. Metabias tests supported these findings as results were all non-significant (all *p’s* > 0.58). No significant clusters/peaks survived correction for the Cocaine > Heroin contrast. No study had an *I^2^* value that exceeded 32% suggesting that the between-study variance was low with most variance attributed to within study sampling error.

**Figure 5.**
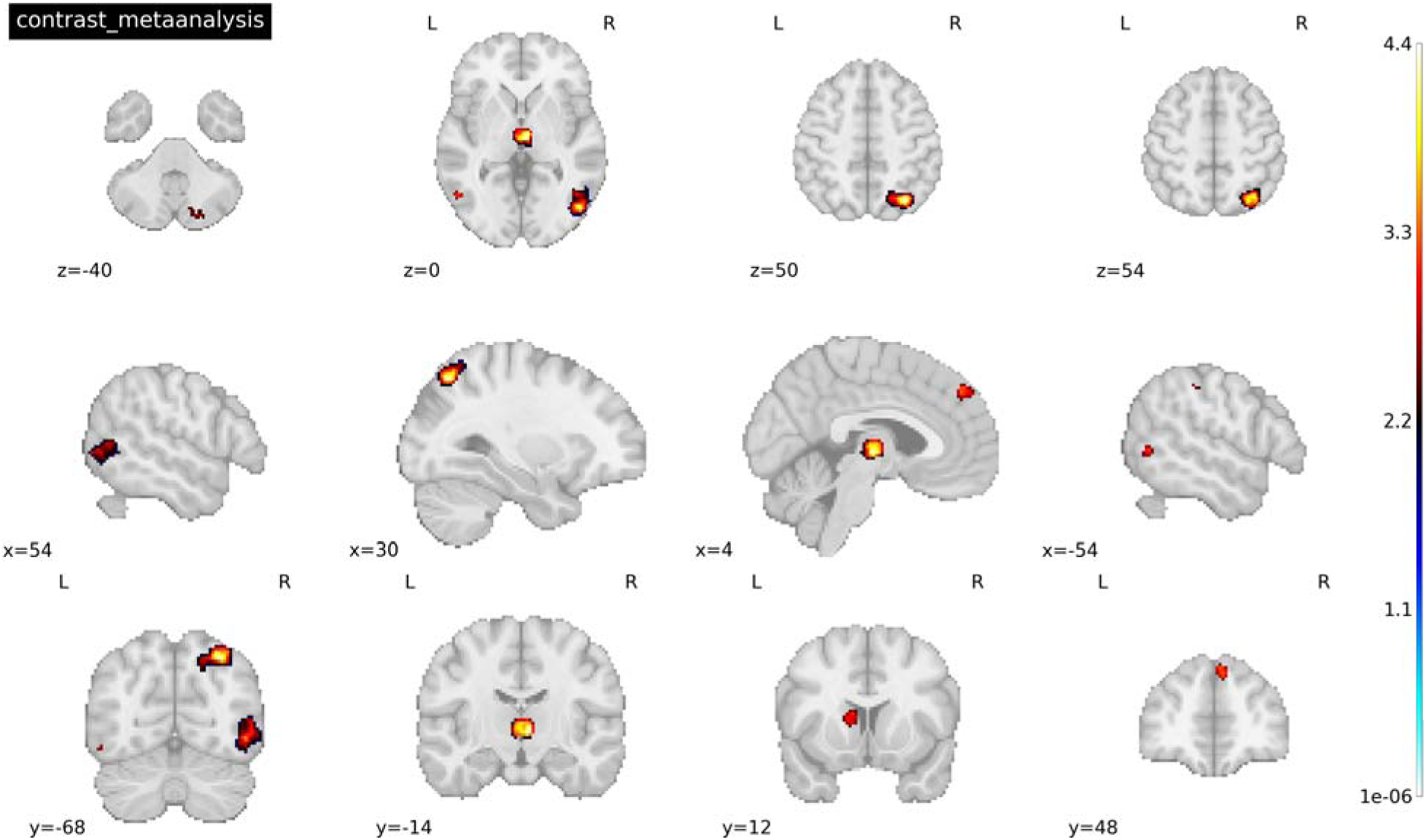
Peak activations associated with the contrast analysis of heroin > cocaine subgroups for drug > neutral cues. Threshold was set at p<.005 TFCE-FWER and cluster size ≥ 10 voxels. Coordinates reported in MNI space.

**Table 6.** Location of neural activations for Heroin > Cocaine contrast of drug > neutral cue studies. Peaks arranged by decreasing voxel size.

| ID | Region | X | Y | Z | Voxels | P | SDM - Z | Bias Test (P value) | $I^2$ |
| --- | --- | --- | --- | --- | --- | --- | --- | --- | --- |
| 1 | Right inferior temporal gyrus, BA 19 | 48 | -74 | -4 | 628 | <0.001 | 4.313 | 0.603 | 32.199 |
| 2 | Right superior parietal gyrus, BA 7 | 30 | -68 | 50 | 473 | <0.001 | 4.387 | 0.598 | 11.118 |
| 3 | Right anterior thalamic projections | 6 | -14 | 2 | 297 | <0.001 | 4.345 | 0.641 | 7.040 |
| 4 | Left caudate nucleus | -10 | 10 | 10 | 94 | 0.01 | 3.376 | 0.716 | 6.025 |
| 5 | Right superior frontal gyrus, BA 9 | 6 | 48 | 40 | 81 | 0.01 | 3.318 | 0.703 | 16.626 |
| 6 | Right cerebellum, crus II | 20 | -80 | -40 | 67 | 0.03 | 2.655 | 0.771 | 0.739 |
| 7 | Left middle temporal gyrus, BA 37 | -54 | -62 | 2 | 54 | 0.02 | 3.169 | 0.712 | 4.254 |
| 8 | Left inferior parietal (excluding supramarginal and angular) gyri, BA 2 | -50 | -32 | 46 | 26 | 0.03 | 2.821 | 0.720 | 8.739 |

### 3.5 Exploratory Meta-Regression

An important consideration when examining activation differences across groups is whether they are moderated by the lifetime length of use of the substance, as resulting activation patterns to drug cues may vary in accordance with acute and chronic usage. To address this question, we conducted an exploratory meta-regression examining a whole-brain interaction between drug type and average length of use (in months). Fifteen of the eighteen original studies were used for this analysis, as three studies did not provide average length of use data (see Table 1 for full list of included studies). This exploratory analysis revealed a negative interaction in clusters including the left superior frontal gyrus (both the medial (-2,62, 20) and dorsolateral areas, (- 18.38,46), which suggests that activation here decreased as length of use increased in people who use heroin (see Table 7 and Figure 6). Low between-study heterogeneity was found for each significant peak (*I^2^* = 1.48-3.14%). Funnel plots were symmetric, suggesting that none of the results were a function of a small subset of included studies or by studies with a small sample size. The potential publication bias tests were all non-significant (p = .998). No significant clusters of activation survived whole-brain correction for people who use cocaine. No study had an *I^2^* value that exceeded 32%.

**Figure 6.**
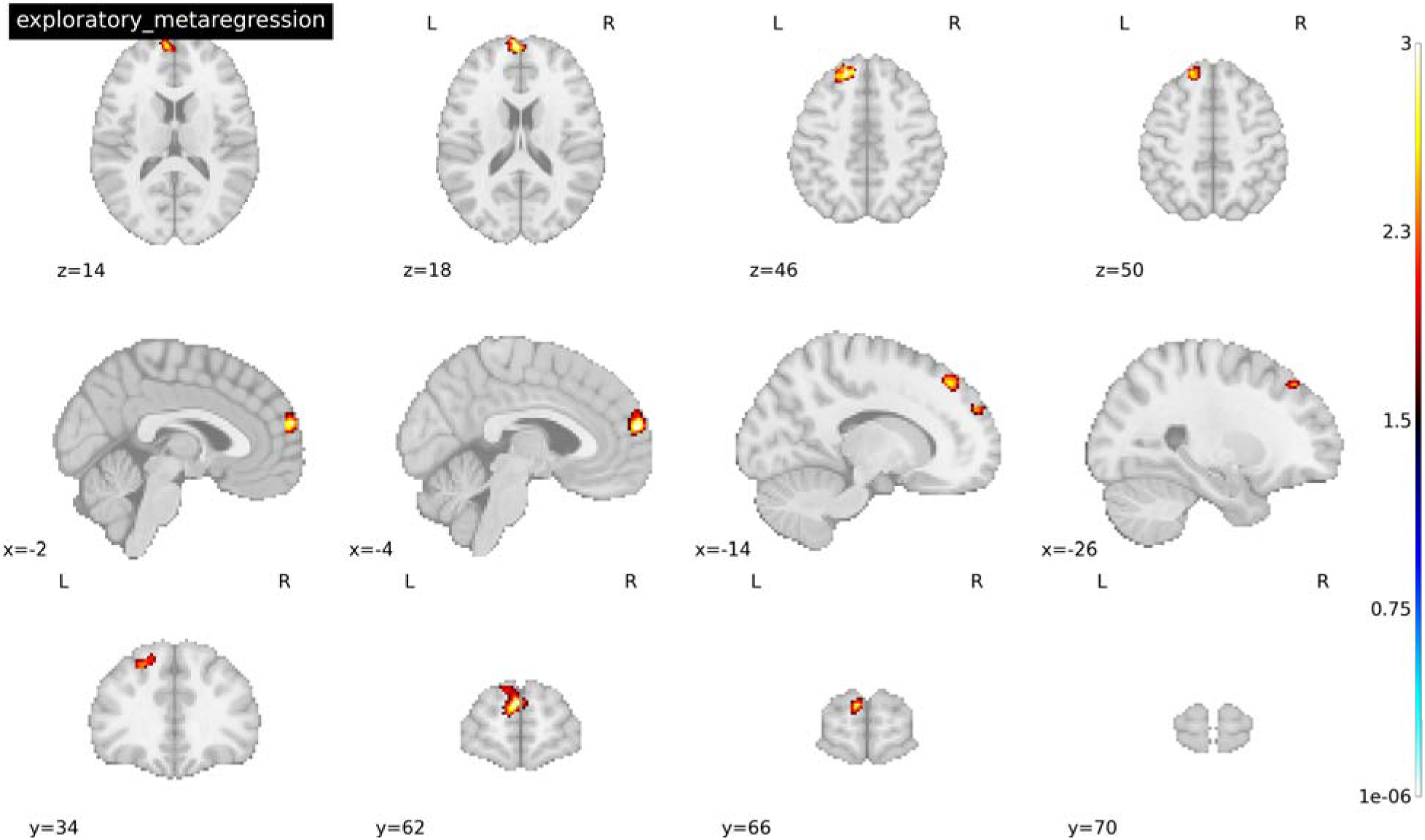
Peak activations across 15 studies of the interaction between length of use and drug type for drug > neutral cues. Threshold was set at p<.005 TFCE-FWER and cluster size ≥ 10 voxels. Coordinates reported in MNI space.

**Table 7.** Location of neural activations for exploratory meta-regression of interaction of drug (Heroin and Cocaine) for drug > neutral cues and average length of use. Peaks arranged by decreasing voxel size.

| ID | Region | X | Y | Z | Voxels | P | SDM - Z | $I^2$ |
| --- | --- | --- | --- | --- | --- | --- | --- | --- |
| 1 | Left superior frontal gyrus, medial, BA 10 | -2 | 62 | 20 | 271 | <.001 | -3.017 | 1.48 |
| 2 | Left superior frontal gyrus, dorsolateral, BA 9 | -18 | 38 | 46 | 193 | <.001 | -3.003 | 3.15 |

## 4. Discussion

In the present study, we sought to investigate the differential neural underpinnings of these two major drug classes with respect to drug cue reactivity, a clinically significant process associated with recurrent use. We specifically focused on studies implementing cue-reactivity paradigms, a gold-standard approach to the clinical neurobiology of craving, with a focus on responses to drug cues relative to neutral cues. Our results demonstrate that there are previously underappreciated differences in activation between people who use heroin and people who use cocaine. Specifically, increased (rather than decreased) activation in dopaminergic targets such as the striatum and mPFC, as well as the temporal and parietal gyri and cerebellum were found in people who use heroin that are not readily attributable to demographic or clinical differences across groups (See Tables 2 & 3).

### 4.1 Aggregate and Substance Specific Effects

Aggregate mean analysis results revealed significant activation in the posterior cingulate gyrus, a region within the default mode network (DMN). The posterior cingulate, as well as the DMN at large, have been (e.g., right posterior cingulate gyrus) widely implicated in cue-reactivity, as well as other paradigms, across substance use disorders (Englemann et al., 2012; Janes et al., 2016; Devoto et al., 2020; Wang et al., 2020). Further, our results also revealed significant activation in the superior and inferior frontal gyrus, which converge with recent meta-analyses, though not all specific to cue-reactivity (Chase et al., 2011; Hill-Bowen et al., 2021; Klugah-Brown et al., 2020). In sum, our results are in line with previous findings as well as behavioral models of addiction suggesting dysfunction in habit development, modulation of behavior, and cognitive control (Buhle et al., 2014; Klugah-Brown et al., 2020; Everett & Robbins, 2016; Koob & Volkow, 2016; Wiers et al., 2015; Zilverstand et al., 2018).

Results from the mean analysis of heroin studies revealed significant activation in the left cingulate, the right superior and inferior frontal gyrus, the right anterior thalamic projections, the right and left amygdala, the left caudate nucleus, and the right cerebellum. These findings are consistent with rodent work indicating that acute heroin administration within the VTA enhanced calcium signaling in dopaminergic neurons (as well as in serotonergic neurons within the dorsal raphe nucleus (DRN) (Wei et al., 2018), which in turn was associated with increased locomotor function. We note that similar patterns of results were reported in this study for administration of cocaine into the VTA and DRN (Wei et al., 2018), though we did not see consistent recruitment of dopaminergic regions in our mean analysis of cocaine studies. Instead, results for the cocaine mean analyses revealed significant peaks of activation in the right precentral gyrus. However, the influence of heterogeneity on cocaine results could not be rigorously tested due to a lack of data of clinical features. Our findings converge with some prior fMRI studies examining cue-reactivity in people who use heroin and people who use cocaine. For example, Yang et al. (2009) conducted a study examining neural responses to cue-reactivity in people who use heroin, reporting activation in the amygdala and cerebellum. Studies have also implicated increased recruitment of frontal regions and reward circuity, albeit with some variation in the specific peak locations when compared with our findings (Yang et al., 2009; Pollard et al., 2023). However, support for this result is variable, with other studies finding decreased activation in the left precentral gyrus (Moeller et al., 2010). It is difficult to compare these studies as methodology and task design varies greatly, however, indicating a need for replication of these findings using consistent task designs.

### 4.2 Between Substance Effects

Interestingly, and contrary to our hypotheses, our direct comparison of cue reactivity between substances revealed increased recruitment of dopaminergic regions and other areas in people who use heroin relative to people who use cocaine. Given the robust literature on the involvement of reward processing regions such as the striatum and PFC across cocaine, heroin and other substances (Yan et al., 2023; Klugah-Brown et al., 2020; Hobkirk et al., 2019; Cooper et al., 2019; Motzkin et al., 2014; Patel et al., 2013; Volkow et al., 2010; Sell et al., 2001), we did expect to find at least some regions of overlap, but our meta-analysis may have been underpowered to detect such effects. Though, this could be explained by differential effectiveness of drugs eliciting cue reactivity, in other words, heroin cues were more effective at eliciting cue-reactivity. A recent study found that when comparing cue induced and tonic craving between opioid and stimulant users in treatment, opioid cues consistently resulted in greater reports of craving (Hochheimer et al., 2023). Such results may in turn support the increased number of regions recruited in our sample of heroin cue reactivity studies. Alternatively, it is also possible that users of cocaine in the studies in our sample may be better able to engage regulatory mechanisms during their responses to cue-induced craving as suggested by dlPFC activation, a region consistently implicated in regulation of craving (George & Koob, 2013; Hayashi et al., 2013; Li et al., 2017; Brevers et al., 2023; Kober et al., 2010).

However, given the known neuropharmacological and clinical differences between these substances, the divergence in neural activation that emerged in our study is plausible. Support for this possibility comes from animal models that have demonstrated distinct recruitment of subpopulations of neurons within the striatum and PFC when comparing neural responses to cocaine and heroin (Ettenberg et al., 1982; Pettit et al., 1984; Chang et al., 1998; Baldani et al., 2011). Further, in spite of differing locations of activation associated with to heroin and cocaine cues, there is evidence from other domains to suggest that differing locations of activation can be mapped back to the same network, ultimately implicating an overall common pathway (Padmanabhan et al., 2019; Stubbs et al., 2023; Jousta et al., 2022). Such a pattern suggests that viewing all substances as having the same neurobiological substrates because of common activation of regions such as the striatum and PFC, to some degree may be an It is therefore plausible that the activation differences we observed reflect preliminary neural underpinnings for the cognitive, behavioral, and clinical differences between heroin and cocaine use, including frequency of use and impulsive behavior, but replication and further research is needed (Hser et al., 2008a; Hser et al., 2008b; Lejuez et al., 2005; Bornovalova et al., 2005).

For example, cocaine and heroin have both been implicated in attentional dysfunction though the way in which each drug disrupts attention varies whereby individuals who use heroin displayed delayed attentional responses, whereas user of cocaine demonstrated increased interference with attention (Bjork et al., 2022). Opioid use has also been linked to impacts on decision-making including decreased response inhibition and delayed gratification (Psederska & Vassileva, 2023). Opiate use may affect other aspects of decision making, such as risky choices, differentially depending on length of abstinence (Psederska & Vassileva, 2023). Our results may parallel these findings, as both cocaine and heroin users show general activation in regions associated with attention and cognitive control (e.g., prefrontal cortex, parietal regions) however, users of heroin seem to show more significant activation in regions associated with cognitive control compared to users of cocaine, a finding which may related to the differences in attentional disruptions found in behavioral tasks (Bjork et al., 2022; Psederska & Vassileva, 2023).

A recent study comparing whole-brain white matter abnormalities in users of both substances found that while both people who use heroin and people who use cocaine showed abnormalities in white matter, people who use heroin showed significantly decreased fractional anisotropy overall in the brain compared with people who use cocaine (Gaudreault et al., 2022), suggestive of decreased white matter integrity. Our results may dovetail with these in that we find studies of people who use heroin showing more diffuse activation than people who use cocaine in dopaminergic regions, specifically the right inferior and left middle temporal gyrus, the right thalamus, and the right cerebellum. Additional evidence from studies examining volumetric impacts of cocaine and heroin use have also converged on the involvement of the nucleus accumbens, which has shown significant gray matter reduction, in people who use both substances when compared to healthy controls; however, we note that this pattern was more robust in people who use heroin (Ceceli et al., 2022; Seifert et al., 2015; Carlezon & Thomas, 2009). Similar findings were reported in the vmPFC (Ceceli et al., 2022).

These observed volumetric differences may suggest biomarkers of susceptibility to substance (ab)use or consequences of prolonged use, which may in turn be related to alterations in functional activation (Ceceli et al., 2022; Seifert et al., 2015). The incorporation of both functional and volumetric measurements, while controlling for contextual factors, (e.g., duration of use, recency of use, age of onset) when examining the implications of substance use in future studies may help address these questions (Oakes et al., 2007).

### 4.3 Length of Use Moderates Heroin Cue Reactivity Activation

Our exploratory meta-regression demonstrated that activation in the left superior frontal gyrus, both the medial and dorsolateral areas, decreased as length of use increased in people who use heroin, controlling for age. However, no significant relationship between cocaine and length of use was found. Literature examining the effect of duration of use on brain activation is limited; however, our findings fit within literature suggesting that drug use is known to impact the prefrontal cortex (Goldstein & Volkow, 2012). A model put forth by Volkow et al. (2010) posits that one of the effects of chronic drug use is decreased reward sensitivity or hypoactivation in the prefrontal cortex. It is unclear whether this is a direct reflection of the effects of chronic drug use on the brain or perhaps better explained by the tolerance hypothesis––i.e., that over time, one needs an increasing amount of a drug to produce the same effect (Peper, 2004). Additionally, Ferenzci et al. (2016) also found a similar pattern of deactivation in the PFC with length of substance use, though they also demonstrated a subsequent increase in BOLD activation in the striatum. While our findings did not show an increased BOLD response in the striatum, this relationship warrants more exploration as our lack of findings in this area may be a function of duration of use for study participants and our small sample size. This may require comparison across casual vs chronic use and examining developmental samples in future studies. Further, larger samples could evaluate both length of use and amount of use over time to explore this relationship further. Additionally, future studies should further examine the relationship between length of use and heroin as our study may not have incorporated enough variation in length of cocaine use to fully capture whether a relationship was meaningful.

### 4.4 Limitations

Limitations of this study include a small sample size due to limited availability of literature for within-group cue-reactivity studies examining only people who use heroin or cocaine. The small sample size impacts the generalizability of these results to widespread populations of people who use cocaine and heroin. Another limitation of the current study is the lack of or inconsistencies in demographic and clinical data collected across studies further limiting full consideration of these variables as moderators. Important co-variates such as additional/co-occurring diagnoses, amount of use, frequency of use, etc. need to be collected across all studies to enhance generalizability and provide opportunities for answering more complex and nuanced questions related to drug addiction. Further, there was variability across studies in the duration of use and time since recent use, preventing the ability to control for these factors in the analysis.

### 4.5 Future Considerations

Given the clinical and behavioral findings that social context is related to drug choice, future studies should consider incorporating a social context into cue-reactivity tasks. This would expand the clinical and behavioral findings to include the effects of social context on functional activation. Future studies should consider implementing a standardized demographic and clinical data collection procedure that includes relevant contextual factors (e.g., time since last use, length of use, amount used) to enhance external validity and better understand the relationship between these factors and neural responses to drug cues (Ekhatiari et al., 2022). Further, future research should examine how the nuanced activation differences identified here may relate to differences identified in other areas of substance use, such as responsiveness to treatment (Lichenstein et al., 2021). Lastly, future longitudinal studies may help answer whether and how such neural activation patterns during cue reactivity precede or succeed problematic substance use.

Taken together, our findings show that while there is some overlap in neural activation across heroin and cocaine use, there are distinct neural patterns uniquely associated with each substance. Additionally, these differential activation patterns in people who use heroin may be a function of length of use. Therefore, it is imperative that research continue to evaluate both the convergence and divergence of activation patterns across substances, as well as individual differences in length of use among other variables in order to relate such findings to behavioral differences as well as individualized treatments.

## Data Availability

All fMRI coordinates included in the meta-analysis reported in this manuscript can be found on the Open Science Framework (https://osf.io/gmu5v/).

## CredIT Statement

Jordan Dejoie (Data Acquisition, Methodology, Writing – original draft, Visualization). Nicole Senia (Conceptualization, Data Curation), David Smith (Conceptualization, Methodology, Writing – review & editing, Visualization), Anna Konova (Conceptualization, Writing – review & editing, Visualization), Dominic Fareri (Supervision, Formal Analysis, Writing – review & editing, Visualization)

## Data Availability

All text files used in this meta-analysis are accessible via the Open Science Framework at https://osf.io/gmu5v.

## Declaration of Competing Interests

The authors hereby declare that there are no competing financial or personal interests that could have appeared to influence the work reported in this paper.

## Ethics Statement

Not applicable due to meta-analytic study.

## Notes

### Competing Interest Statement

The authors have declared no competing interest.

### Funding Statement

This manuscript is supported in part by funding from the National Institute on Mental Health (R15MH122927 to DSF), the National Institute on Drug Abuse (R01DA053282, R01DA054201 to ABK) and the National Institute on Aging (RF1-AG067011 to DVS).

### Author Declarations

This meta-analysis only used coordinates reported from contrast analyses in published fMRI studies of cue reactivity in users of cocaine and heroin. All data input into this meta-analysis were from previously published papers found via PubMed and Google Scholar. No identifying information or raw data from any individual was used in this manuscript. All studies that were used are included in a table in the manuscript and in the reference section of the manuscript. All fMRI coordinates from relevant analyses can be found on the Open Science Framework (https://osf.io/gmu5v/).

### Summary of Updates

Revisions in this version of the manuscript focus on: increasing readability and conciseness throughout the manuscript; increasing methodological specificity; enhancing visualizations of all figures; reporting results of permutation analyses with a larger number of permutations.

